# On the Front (Phone) Lines: Results of a COVID-19 Hotline in Northeast Ohio

**DOI:** 10.1101/2020.05.08.20095745

**Authors:** David Margolius, Mary Hennekes, Jimmy Yao, Douglas Einstadter, Douglas Gunzler, Nabil Chehade, Ashwini R. Sehgal, Yasir Tarabichi, Adam T. Perzynski

## Abstract

**Importance:** Severe acute respiratory syndrome coronavirus (SARS-CoV-2) and the associated coronavirus disease of 2019 (COVID-19) have presented immense challenges for health care systems. Many regions have struggled to adapt to disruptions to health care practice and employ systems that effectively manage the demand for services.

**Objective:** To examine the effectiveness of the first five weeks’ of a 24/7 physician-staffed COVID-19 hotline.

**Design:** Cohort study using electronic health records.

**Setting:** A single large health care system in Northeast Ohio.

**Participants:** During 5 weeks of operation, 10,112 patients called the hotline (callers) and were evaluated by a registered nurse (RN) using standardized protocols. Of these, 4,213 (42%) were referred for a physician telehealth visit (telehealth patients). The mean age of callers was 42 years. 67% were female, 51% white, and 46% were on Medicaid or uninsured.

**Intervention:** Physician telehealth visits for COVID-19.

**Main Outcomes and Measures:** We describe clinical diagnosis, patient characteristics (age, sex race/ethnicity, smoking status, insurance status), and visit disposition. We use logistic regression to evaluate associations between patient characteristics, visit disposition and subsequent emergency department use, hospitalization, and SARS-Cov-2 PCR testing.

**Results:** Common caller concerns included cough, fever, and shortness of breath. Most telehealth patients (79%) were advised to self-isolate at home, 14% were determined to be unlikely to have COVID-19, 3% were advised to seek emergency care, and 4% had miscellaneous other dispositions. A total of 287 (7%) patients had a subsequent ED visit, and 44 (1%) were hospitalized with a COVID-19 diagnosis. Of the callers, 482 (5%) had a COVID-19 test reported with 69 (14%) testing positive. Among patients advised to stay at home, 83% had no further face-to-face visits. In multivariable results, only a physician recommendation to seek emergency care was associated with emergency room use (OR=4.73, 95%CI 1.37-16.39, p=.014). Only older age was associated with having a positive test result.

**Conclusions and Relevance:** Robust, physician-directed telehealth services can meet a wide range of needs during the acute phase of a pandemic, conserving scarce resources such as personal protective equipment and testing supplies and preventing the spread of infections to patients and health care workers.

**Key Points:** *Question:* What is the feasibility and effectiveness of physician telehealth services during a pandemic?

*Findings:* In this cohort study of a COVID-19 telehealth hotline that included 10,112 callers and 4,213 physician telehealth visits, most patients (79%) were advised to self-isolate at home, 14% were found unlikely to have COVID-19, 4% dispositions (e.g. testing or office visit) and 3% were advised to immediately seek care emergency department. 83% of patients who were advised to stay home did not require in-person visits.

*Meaning:* Physician-directed telehealth services conserve scarce resources and provide effective, equitable care during a pandemic without compromising patient safety.

## Introduction

The coronavirus disease of 2019 (COVID-19) is caused by a novel pathogen (SARS-CoV-2) first detected in Wuhan, China in December 2019.[1] This pathogen has since spread worldwide with >3 million infected and >900,000 cases and >50,000 deaths in the United States[2] before the end of April 2020. Although only a minority of patients with COVID-19 develop severe disease [3], researchers and clinicians have struggled to adapt to the cascade of disruptions to health care practice and to employ systems that effectively manage the demand for services. Local, national and international policymakers and health system leaders around the globe are urgently in need of evidence to guide decision making on effective clinical care strategies as a component of pandemic response.

During the 2009 H1N1 pandemic, nurse triage lines reduced in-person visits for influenza-like symptoms, were cost effective, had a high degree of satisfaction among callers, and were able to reach rural and uninsured populations.[4,5,6,7] Reaching vulnerable populations and those of lower socioeconomic status (SES) during a pandemic is of particular importance as individuals of lower SES have greater barriers to information access and are more likely to adopt incorrect protective behaviors.[8]

To further promote telehealth in the COVID-19 era, throughout March and April of 2020 federal and state policies have been altered to remove or ease geographic restrictions and other regulatory barriers to telehealth visits.[10] Telehealth is well-suited for use during pandemics as clinicians can provide care and consultation to isolated populations and counter the surge in demand for medical care while using telehealth as a form of electronic personal protective equipment.[9, 11, 12, 13] Telehealth in the COVID19 era is, thus, theorized as a useful strategy in “forward triage,” the sorting of patients prior to their arrival in the emergency department[13,14] and as a means to provide patient guidance and reassurance[11].

On Monday, March 9^th^, the first 3 cases of coronavirus disease 2019 (COVID-19) were diagnosed in Northeast Ohio.[15] On March 13^th^, our health care system launched a RN triage-linked, 24-hour availability, physician-staffed hotline to assess, advise and treat individuals who called with symptoms that could be COVID-19 related. Over the next five weeks, the RN triage line received more than 12,000 calls resulting in more than 5,000 physician telehealth visits. While other institutions have reported on the establishment of triage system protocols within their electronic health records (EHRs) and have updated patient portals to provide patients with self-triage and self-scheduling abilities[15-17], to our knowledge this is the first study to examine use of a COVID-19 hotline that provides patients with direct physician care.

## Methods

### Overview

This is a cohort study of patients who called a COVID-19 hotline at MetroHealth, a large urban safety-net health care system in Cuyahoga County in Northeast Ohio. The study was approved by the MetroHealth Institutional Review Board.

### Setting

This study took place at an academic health care system in Cleveland, Ohio and surrounding municipalities. The system employs more than 7800 individuals and consists of four hospitals, four emergency departments, and twenty health centers. In 2019, the system delivered care to more than 298,000 unique patients. According to the Ohio Department of Health, Cuyahoga County has among the highest number of COVID-19 cases among Ohio counties and the highest number of deaths. Cuyahoga County (including the City of Cleveland, Ohio) is also among the most densely populated and socioeconomically disadvantaged communities in the United States.[18]

### Intervention

All individuals in Northeast Ohio with questions or concerns about COVID-19 were invited to call a dedicated hotline if they had questions or concerns about COVID-19 (Figure 1). Callers are prompted to press 1 to hear information about COVID-19 or press 2 to speak with a nurse about questions or signs and symptoms of COVID-19. Using standard protocols based on the caller’s symptoms, the nurse determined if further evaluation for COVID-19 was warranted and, if so, scheduled the patient for a same-day telehealth visit with a physician. A total of 91 physicians from family medicine, medicine-pediatrics, general internal medicine, pediatrics, and otolaryngology have staffed the hotline. Physicians evaluate the patient’s condition over the phone and recommend a treatment plan. Patients who completed a physician telehealth visit received a follow-up call from a care coordinator within 24 hours to assess for any change in symptoms. During the follow-up call, the care coordinator also assessed the patient for limitations in basic living needs and offered to connect them with services provided by the MetroHealth Institute for H.O.P.E. (Health, Opportunities, Partnerships, Empowerment), including home delivery of food and prescriptions, behavioral health visits, and spiritual care.

**FIGURE 1:**
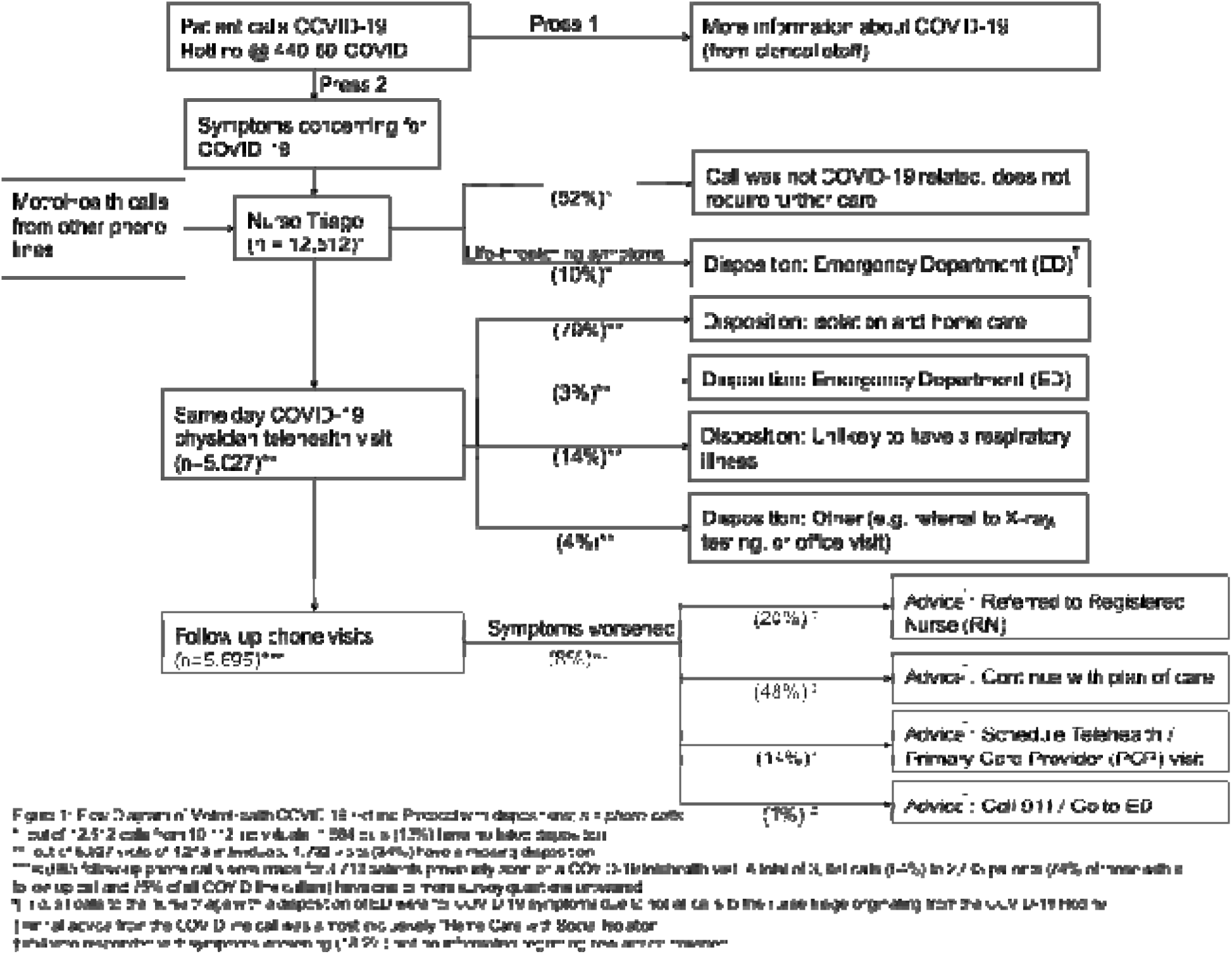
COVID-19 Hotline Patient Flow

### Measures

We queried the electronic health record (Epic^TM^, Verona, WI) to extract data on patients who called the COVID-19 hotline and completed nurse triage only or were referred for a telehealth visit with a physician. We collected patient characteristics (age, sex, race/ethnicity, insurance type, smoking status) and clinical variables directly relevant to understanding the social epidemiology of the COVID-19 hotline (symptom protocols, visit disposition, visit diagnoses). We also analyzed data from follow-up calls with regards to changes in health status and self-described basic needs. Race and ethnicity categories included Non-Hispanic White, Non-Hispanic Black, Hispanic, and other. Insurance type categories included Medicaid, Medicare, Commercial, Employee Insurance (a form of commercial insurance covering The MetroHealth System employees), and Uninsured. Smoking status was defined as current, former, or never/unknown.

### Outcomes

We evaluated four outcomes: 1) emergency room visit likely related to COVID-19 subsequent to hotline telehealth visit, (2) hospitalization due to COVID-19 subsequent to hotline telehealth visit, (3) SARSCoV-2 PCR test ordered subsequent to telehealth visit, and (4) positive SARS-CoV-2 PCR test subsequent to telehealth visit.

### Statistical Analyses

Patient disposition following physician visits was reported descriptively in three categories – patient advised on COVID-19 isolation and home care, patient advised to visit the emergency department, or other (unlikely to have a viral respiratory illness or illness related to COVID-19). Visit disposition and patient characteristics were then incorporated as explanatory variables into a series of multivariable logistic regression models for each of the four outcomes described above. The models were used to calculate odds ratios and 95% confidence intervals to measure the association between each explanatory variable after adjusting for other model covariates and each outcome of interest. Regression analyses were conducted using SAS Version 9.4 (Cary, NC). Missing data was handled with listwise deletion. All statistical tests are two-tailed with significance defined as p < .05.

## Results

The complete study flow and most common visit dispositions are represented in Figure 1. Between March 13 and April 20, 2020, there were a total of 12,512 calls to the RN triage line from 10,112 unique individuals. A total of 5,027 RN triage calls from 4,213 patients were referred for a telehealth physician visit. An additional 96 physician telehealth visits had no preceding RN triage call. Demographic characteristics of the nurse triage-only and the physician COVID-19 line patients are presented in Table 1. Daily call volume was steady throughout the five-week study period and the cumulative increase continued almost parallel to growth in confirmed COVID-19 cases for Cuyahoga County (Figure 2). For all calls, common reasons for the call were cough (22.3%), advice/health education (15.6%), difficulty breathing (6.1%), fever (4.7%), and flu-like illness (4.0%). Similarly, the most common RN protocols used were for cough (11.0%), chest pain (4.8), sore throat (4.0%), abdominal pain (3.4%), and fever (3.3%). Among all calls to the RN triage line, 38% were referred for a same-day physician telehealth visit. 10% were advised to go immediately to the emergency room, and 52% were not COVID19-related or required no additional care. Of the hotline callers, 482 (5%) had one or more COVID tests reported with 69 (14%) testing positive. A total of 287 (7%) telehealth patients had a subsequent ED visit, and 44 (1%) were hospitalized with a COVID-19 diagnosis. Among those patients advised to stay at home at their first physician hotline visit, 83% recorded no further face-to-face clinical encounters.

**TABLE 1:** Descriptive characteristics of COVID-19 Hotline Patients.

| **TABLE 1: Descriptive characteristics of COVID-19 Hotline Patients.** |  |  |  |
| --- | --- | --- | --- |
| **Characteristic** | **All** | **RN Triage Line only** | **COVID-19 MD Line** |
| Individuals | 10,208 | 5,995 | 4,213 |
| Mean Age, years | 41.9 | 41.9 | 42.0 |
| Female, % | 67.4 | 66.8 | 68.3 |
| Smoking Status, % |  |  |  |
| Current Smoker | 20.3 | 20.3 | 20.3 |
| Former Smoker | 28.0 | 29.3 | 26.1 |
| Never smoker | 44.8 | 44.5 | 45.3 |
| Unknown / Not Asked | 6.9 | 5.9 | 8.3 |
| Race, % |  |  |  |
| White | 51.2 | 47.8 | 56.0 |
| Black | 38.0 | 41.9 | 32.5 |
| Hispanic | 5.3 | 5.4 | 5.3 |
| Other / Unknown | 5.5 | 4.9 | 6.2 |
| Insurance, % |  |  |  |
| Commercial | 37.0 | 31.7 | 44.1 |
| Medicare | 16.7 | 19.5 | 12.9 |
| Medicaid | 35.2 | 39.3 | 29.6 |
| Uninsured | 10.6 | 8.7 | 13.1 |
| Other | 0.6 | 0.8 | 0.3 |
| * Note: 96 patients had a COVID line call without a prior RN Triage call |  |  |  |

**FIGURE 2:**
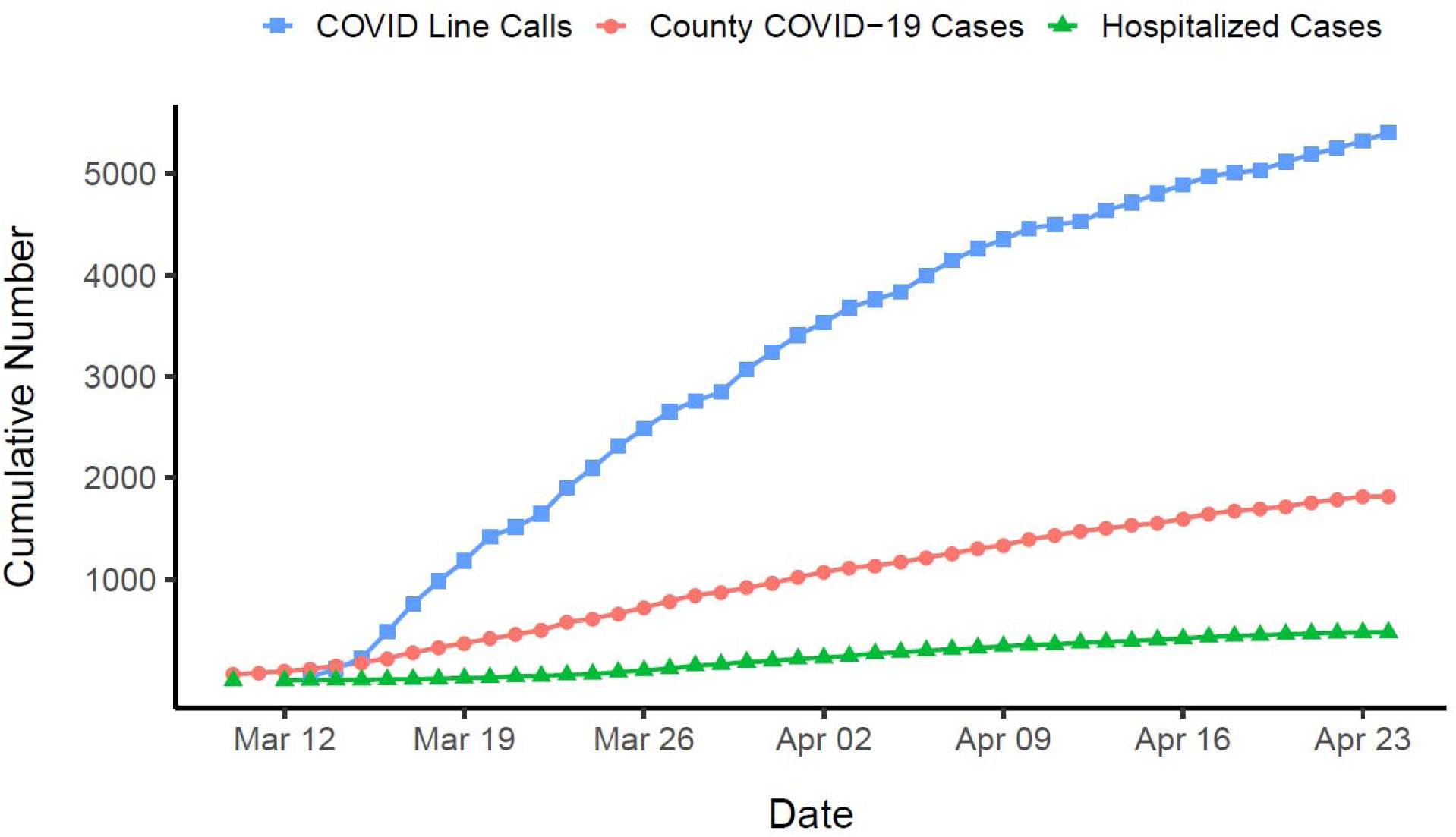
COVID-19 Cases (Hotline and Cuyahoga County) and Hospitalizations (County)

For the patients with a physician phone visit, common visit diagnoses included respiratory symptoms (43.0%), URI unspecified type (5.7%), cough (5.5%), viral URI with cough (3.6%), and sore throat (2.1%). Less than 2% of scheduled phone calls were “no show.” Among all physician telephone visits, 79% were advised to stay at home, self-isolate, and continue monitoring symptoms. In 14% of visits, physicians felt the patient was unlikely to have a COVID-related problem. 5% were advised to go to the emergency department or seek additional non-emergency care or testing.

Of the 4,213 patients with a physician phone visit, 3,713 (88%) received one or more care-coordinator follow-up calls. At follow-up, 92% of patients reported their symptoms had not worsened and 67% reported having connected with the follow-up recommended on the initial physician phone visit. Some patients (16%) reported feeling overwhelmed, anxious, or isolated due to COVID-19, and 3% reported needing help to manage their basic needs. Those requiring help with basic needs were offered services from the MetroHealth Institute for H.O.P.E., which has provided these patients with multiple services including food deliveries, prescription deliveries, social work support, behavioral health telephone visits, and faith-based comfort calls from pastoral care personnel.

Findings from multivariable logistic regression models including patient demographics, smoking status and physician telephone disposition are presented in Table 2. Emergency department visits and hospitalizations were positively associated with a telehealth visit disposition indicating that the patient was high risk and should seek emergency care. Receipt of COVID-19 testing was associated with older age, current smoking status, employee health insurance, and a telephone visit disposition of high risk or self-isolation. Finally, among those who received testing, a positive test result was associated with older age.

**TABLE 2:** Logistic Regression Results for Patient Characteristics Associated with (1) ED Visit (2) COVID-19 Hospitalization (3) SARS-CoV-2 PCR Testing and (4) Positive SARS-CoV-2 PCR Test

|  | ED Visit<br>N=2569, c=0.65 |  |  | Hospitalization<br>N=2569, c=0.77 |  |  | SARS-CoV-2 Test<br>N=2569, c=0.76 |  |  | Positive SARS-CoV-2<br>N=249, c=0.67 |  |  |
| --- | --- | --- | --- | --- | --- | --- | --- | --- | --- | --- | --- | --- |
| *Parameter* | *Odds ratio* | *95% CI* | *p* | *Odds ratio* | *95% CI* | *p* | *Odds ratio* | *95% CI* | *p* | *Odds ratio* | *95% CI* | *p* |
| Age (per 10 years) | 1.14 | 0.95-1.37 | 0.15 | 1.09 | 0.77-1.54 | 0.62 | 1.13 | 1.03-1.24 | 0.01 | 1.47 | 1.10-1.96 | 0.01 |
| Sex (Female) |  |  |  |  |  |  |  |  |  |  |  |  |
| Male | 1.29 | 0.72-2.22 | 0.42 | 0.70 | 0.22-2.25 | 0.55 | 1.20 | 0.88-1.62 | 0.24 | 1.40 | 0.68-2.93 | 0.36 |
| Race (NH White) |  |  |  |  |  |  |  |  |  |  |  |  |
| NH Black | 1.03 | 0.56-1.87 | 0.93 | 0.85 | 0.28-2.60 | 0.77 | 0.84 | 0.61-1.15 | 0.28 | 1.68 | 0.78-3.62 | 0.19 |
| Hispanic | 1.57 | 0.59-4.19 | 0.36 | 1.19 | 0.14-9.80 | 0.87 | 0.61 | 0.30-1.27 | 0.19 | 0.68 | 0.08-6.01 | 0.73 |
| Other | 0.93 | 0.28-3.14 | 0.90 | 1.17 | 0.14-9.67 | 0.88 | 1.28 | 0.75-2.17 | 0.36 | 0.66 | 0.16-2.66 | 0.56 |
| Insurance type (Commercial) |  |  |  |  |  |  |  |  |  |  |  |  |
| Medicaid | 0.86 | 0.38-1.95 | 0.72 | 2.71 | 0.27-27.24 | 0.40 | 0.97 | 0.60-1.56 | 0.90 | 2.62 | 0.79-8.74 | 0.12 |
| Medicare | 1.18 | 0.47-3.00 | 0.72 | 4.98 | 0.48-51.63 | 0.18 | 1.11 | 0.63-1.94 | 0.72 | 0.56 | 0.12-2.61 | 0.46 |
| Employee | 1.55 | 0.69-3.46 | 0.29 | 11.45 | 1.35-96.95 | 0.02 | 7.13 | 4.82-10.54 | <0.01 | 1.35 | 0.49-3.75 | 0.56 |
| Uninsured | 1.76 | 0.74-4.22 | 0.20 | 4.26 | 0.38-47.79 | 0.23 | 1.15 | 0.68-1.96 | 0.60 | 1.55 | 0.38-6.34 | 0.54 |
| COVID-19 line disposition (Other) |  |  |  |  |  |  |  |  |  |  |  |  |
| Stay at home | 1.95 | 0.91-4.21 | 0.09 | 0.77 | 0.23-2.54 | 0.66 | 1.90 | 1.29-2.78 | <0.01 | 1.08 | 0.40-2.87 | 0.89 |
| High Risk | 4.73 | 1.37-16.39 | 0.01 | 7.77 | 1.61-37.50 | 0.01 | 8.78 | 4.39-17.53 | <0.01 | 0.48 | 0.07-3.14 | 0.44 |
| Smoking (Never, Unknown) |  |  |  |  |  |  |  |  |  |  |  |  |
| Current | 0.67 | 0.32-1.40 | 0.29 | 0.76 | 0.15-3.78 | 0.73 | 0.41 | 0.26-0.65 | <0.01 | 0.53 | 0.14-2.03 | 0.36 |
| Former | 0.70 | 0.36-1.36 | 0.29 | 1.46 | 0.47-4.56 | 0.52 | 0.65 | 0.46-0.92 | 0.02 | 0.70 | 0.29-1.69 | 0.42 |

## Discussion

In this report, we demonstrate the feasibility and effectiveness of a large-scale physician-staffed hotline in providing care and disseminating information to the general public of a large metropolitan area amidst a global pandemic. Strengths of our study include a large sample consisting of many individuals who were not tested despite concerns about having COVID-19, care process data from standardized protocols, and outcome data collected from the electronic health records from multiple care systems.

Recent reports have focused on use of a self-administered survey as a mechanism for conserving PPE, citing the difficulty of rapidly creating robust telehealth infrastructure.[21] In contrast, our results indicate that robust, comprehensive, and hospital-integrated telehealth are an effective form of health services during the acute phase of a pandemic and beyond. Telephone hotlines require a minimum of technological capabilities, can be implemented rapidly, and are accessible to those who may not have internet access, especially when facilities with public internet access such as libraries are closed. Past research has indicated that telephone calls yield similar patient health outcomes when compared to video-based appointments [20].

Although some communities have described higher rates of infection, hospitalization, and death among low income and racial and ethnic minority patients, data from our hospital system in Northeast Ohio does not indicate any observed racial or socioeconomic disparities in care process or outcome. Our results indicate that employees of the health care system who called the hotline were more likely to be tested, but this is a function of system policies which referred employees with suspect or proven COVID-19 exposure to call the hotline as well as the need to protect patients from potentially infected care personnel. In our population, we found no association between current smoking status and emergency room visits or hospitalization after a telephone encounter for patients with COVID-19-related symptoms. This finding is consistent with a review of several prior studies outside the United States which found little evidence for the influence of tobacco smoking on COVID-19 outcomes.[22] However, prior investigations and reviews have been confined to case series designs derived from data reported to public health agencies or to hospitalized patients which limit the ability to include important covariates and excludes untested patients reporting acute respiratory illness.[22-25]

The results of our study have several important limitations. We report on a hotline from a single, large health care system serving an urban area of Northeast Ohio. Our findings may not be generalizable to other health systems or areas due to variation in COVID-19 government and public health response. Although we included data available through Epic’s Care Everywhere health information exchange, we may not have captured all data from other regional care systems, and some patients may have received services at other hospitals or emergency rooms that are not available for our analyses. However, two of the three major health systems in Cuyahoga County use the Epic EHR, and COVID-19 test results from all three systems (covering more than 90% of all acute care) were captured in our data, suggesting any missed utilization may be minimal.

Prior research has demonstrated that telephone hotlines are a convenient and efficient mode of information distribution from health care providers to a large population, and our data indicates that a significant number of our study population sought advice or information as the primary reason for calling. [8]. Readily accessible information and medical care during a pandemic is a crucial public health function as it can directly reduce demand for emergency services and efficiently provide large numbers of symptomatic and potentially infected persons guidance about how to stay at home and protect themselves and others. The vast majority of patients with telehealth visits in our population were advised to seek home care, avoiding the need to seek care at a walk-in clinic or the emergency room.

We did not find evidence of disparities by race and ethnicity or insurance type, which leads us to conclude that telephone hotline services are an accessible and equitable form of care delivery during the COVID-19 pandemic. Services that depend on internet access, such as patient portals, have been shown have lower usage among older adults, racial and ethnic minorities and older people, the very populations most affected by COVID-19 severe disease in the United States. Further, results of our multivariable models indicate that primary care physicians assessing patients over the phone make effective decisions about which patients will require use of emergency services and which can be safely managed at home. By transforming a large number of potential in-person visits to telephone visits our study demonstrates that it is possible to conserve increasingly scarce personal protective equipment, appropriately limit testing, and potentially prevent the further spread of infection to patients and health care workers in otherwise busy in-person care settings. DeVoe and colleagues recently published a plea for a regional telehealth primary care extension infrastructure to address the demands being placed on care systems.[28] Our findings provide evidence for the effectiveness of such an approach, and suggest that policymakers and medical and public health leaders should consider widespread implementation of physician-staffed telehealth services as a key component of effective, equitable pandemic response.

## Data Availability

De-identified data can be requested directly from the authors.

## Acknowledgements

Sandra Andrukat, Kim Bauchens RN, MSN, Shari Bolen MD, Karen Cook, Nick Dreher MD, Ryan Johnson

## Funding

The MetroHealth System

## Notes

### Competing Interest Statement

The authors have declared no competing interest.

### Funding Statement

This work was supported by the MetroHealth System. No external funding was involved.

